# Rapid comparative evaluation of SARS-CoV-2 rapid point-of-care antigen tests

**DOI:** 10.1101/2021.07.29.21261314

**Authors:** Anna Denzler, Max L. Jacobs, Victoria Witte, Paul Schnitzler, Claudia M. Denkinger, Michael Knop

**Affiliations:** Center for Molecular Biology of Heidelberg University (ZMBH), Heidelberg, Germany; Department of Infectious Diseases, Virology, Heidelberg University Hospital, Heidelberg, Germany; Department of Infectious Diseases, Division of Tropical Medicine, Heidelberg University Hospital, Heidelberg, Germany; German Cancer Research Center (DKFZ), Heidelberg, Germany; DKFZ-ZMBH Alliance, Heidelberg, Germany

**Keywords:** Comparison of Antigen-detecting rapid diagnostic test, COVID-19, SARS-CoV-2, Testing sensitivity, non-functional AgPOC tests

## Abstract

**Background:** Currently, more than 500 different AgPOCTs for SARS-CoV-2 diagnostics are on sale (July 2021), for many of which no data about sensitivity other than self-acclaimed values by the manufacturers are available. In many cases these do not reflect real-life diagnostic sensitivities. Therefore, manufacturer-independent quality checks of available AgPOCTs are needed, given the potential implications of false-negative results.

**Objective:** The objective of this study was to develop a scalable approach for direct comparison of the analytical sensitivities of commercially available SARS-CoV-2 antigen point-of-care tests (AgPOCTs) in order to rapidly identify poor performing products.

**Methods:** We present a methodology for quick assessment of the sensitivity of SARS-CoV-2 lateral flow test stripes suitable for quality evaluation of many different products. We established reference samples with high, medium and low SARS-CoV-2 viral loads along with a SARS-CoV-2 negative control sample. Test samples were used to semi-quantitatively assess the analytical sensitivities of 32 different commercial AgPOCTs in a head-to-head comparison.

**Results:** Among 32 SARS-CoV-2 AgPOCTs tested, we observe sensitivity differences across a broad range of viral loads (∼7.0^*^10^8^ to ∼1.7^*^10^5^ SARS-CoV-2 genome copies per ml). 23 AgPOCTs detected the Ct25 test sample (∼1.4^*^10^6^ copies/ ml), while only five tests detected the Ct28 test sample (∼1.7^*^10^5^ copies/ ml). In the low range of analytical sensitivity we found three saliva spit tests only delivering positive results for the Ct21 sample (∼2.2^*^10^7^ copies/ ml). Comparison with published data support our AgPOCT ranking. Importantly, we identified an AgPOCT offered in many local drugstores and supermarkets, which did not reliably recognize the sample with highest viral load (Ct16 test sample with ∼7.0^*^10^8^ copies/ ml) leading to serious doubts in its usefulness in SARS-CoV-2 diagnostics.

**Conclusion:** The rapid sensitivity assessment procedure presented here provides useful estimations on the analytical sensitivities of 32 AgPOCTs and identified a widely-spread AgPOCT with concerningly low sensitivity.

## Introduction

In the SARS-CoV-2 pandemic, lateral flow antigen tests were developed as a rapid alternative to SARS-CoV-2 reverse transcriptase quantitative polymerase chain reaction (RT-qPCR)-based diagnostics. Because of their ease of use, lateral flow antigen tests are applicable for point-of-care (POC) and self testing and can therefore be incorporated in the daily life to support viral containment (WHO, Interim guidance, 2020). These tests, in the following referred to as antigen point-of-care tests (AgPOCTs), are meanwhile widely used for SARS-CoV-2 diagnostic and screening purposes. Currently, several hundred different SARS-CoV-2 AgPOCTs brands are commercially available to meet the demand (545 products for professional use are listed by the Federal Institute for Drugs and Medical Devices (Bundesinstitut für Arzneimittel und Medizinprodukte (BfArM)); as of July 27, 2021). However, sensitivity and specificity of the tests are not systematically assessed.

If a test is used by a professional operator, it falls under the ‘low-risk’ category of the European Union directive on In Vitro Diagnostics (IVD), which currently governs marketing authorization for IVDs in Europe. Under this directive, manufacturers can still self-certify COVID-19 tests and waive independent verification of the tests before they are marketed. The validation of the tests, which are offered online and in pharmacies, is therefore not assured in the view of the Paul Ehrlich Institute (PEI), Federal Institute for Vaccines and Biomedical Products, Germany (https://www.pei.de/DE/newsroom/hp-meldungen/2020/200323-covid-19-nat-tests.html;jsessionid=F786872EBB85959AE8DA2B8FCB3ABE00.intranet222?nn=169730). There is also evidence of counterfeiting here. A new legislation governing independent validation by specialized and certified reference laboratories is planned, but will only become effective in March 2022 at the earliest.

If a test is distributed for layperson use, it falls under a ‘higher-risk’ category and requires independent validation. This validation of sensitivity is currently performed by the PEI together with reference laboratories and a list with AgPOCTs passing their criteria is provided (PEI, 2021). AgPOCTs failing the comparative evaluation by PEI will be removed from the list provided by BfArM. This list, however, comprises only products, which were also registered for listing by manufacturers or distributors (https://www.bfarm.de/DE/Medizinprodukte/Aufgaben/Spezialthemen/Antigentests/_node.html), rendering the absence of an AgPOCT from this list difficult to interpret.

Many in-depth AgPOCT characterization studies show that AgPOCT sensitivities can vary substantially. One study reporting on the validation of 122 AgPOCs has recently been published (Scheiblauer *et al*., 2021). The authors found that 26 AgPOCTs do not fulfill the required minimum sensitivity, clearly illustrating that quite a number of circulating AgPOCT are insufficiently sensitive. In addition to this, significant brand-to-brand and lot-to-lot variations were observed (Dinnes *et al*, 2021). These circumstances urge the need for an easy-to-use method to quickly assess AgPOCTs at market entry and periodically thereafter for post-implementation quality control.

In this study, we seeked to establish a procedure to rapidly evaluate a large number of products for their sensitivity, using a small test sample panel and several tests per product. For this we developed a strategy involving pooled samples and four different dilution steps from high to low viral loads, and generated several hundred aliquots thereof. Using this approach we then investigated 32 AgPOCTs, mainly tests currently in use in the local area (Heidelberg, Germany). We compared the results with data from the literature, which enabled us to draw conclusions on the validity of our approach and the performance of the products investigated.

## Methods

### Study design

We tested the analytical sensitivity of a large number of commercially available AgPOCTs by applying pooled samples from nasopharyngeal swabs with defined SARS-CoV-2 viral loads including Ct16, Ct21, Ct25 and Ct28 (∼7.0^*^108 to ∼1.7^*^10^5^ genome copies per ml) as well as a pooled sample obtained from SARS-CoV-2 negative tested persons. Pools were generated using anonymized remnant swab sample material that had been collected for clinical diagnosis of SARS-CoV-2 infection by RT-qPCR carried out by the Center for Infectious Diseases, Virology, Heidelberg University Hospital, Germany. Pharyngeal swab specimens were collected through the nose (nasopharyngeal) and contained in viral transport medium (VTM). Per test, 50 µl of the samples were mixed with the provided lysis buffer of each AgPOCT and the tests were performed strictly according to the manufacturer’s instructions. After the recommended incubation time, images of the test chambers were acquired using a Panasonic Lumix DMC-G70 camera equipped with a Panasonic H-FS12060 objective. AgPOCTs were tested at least in duplicates with the corresponding test samples. Test results were quantified by measuring the background-corrected signal intensities of the test (T) band versus control (C) band in ImageJ (v1.53c) using the “Gels” analysis function usually used for quantification of Western Blot bands. For qualitative evaluation of the visibility of the test bands (positive versus negative score), RGB pictures of AgPOCT results from randomly chosen replicates were evaluated independently by three individuals in a blinded manner. Furthermore, all additional replicates of all AgPOCTs and test samples were scored independently by another person.

### Preparation of test samples from nasopharyngeal swabs

Anonymized, remnant nasopharyngeal swab samples positively and negatively tested for SARS-CoV-2 were obtained between May and July 2021 from the the Center for Infectious Diseases, Virology, Heidelberg University Hospital, Germany. Samples were stored in VTM. The Ct16, Ct21 and negative test samples were prepared by pooling of 12-15 nasopharyngeal swab samples. Cell debris and other solids were removed by centrifugation at 400g for 10min and subsequent transfer of the supernatant into a new tube. Viral RNA was isolated from pools by manual lysis and automated RNA extraction using the QIAamp Viral RNA Mini kit (Qiagen) on a QIAcube Connect device (Qiagen). The cycle threshold (Ct) values of sample pools were determined by RT-qPCR analysis using the LightMix^®^ Modular Sarbecovirus SARS-CoV-2 (TIB Molbiol) with the LightCycler^®^ Multiplex RNA Virus Master (Roche) and LightCycler480 II (Roche). Subsequently, pools were supplemented with 2 % TritonX-100 and c0mplete Ultra protease inhibitor (Roche) and if needed adjusted with dilution buffer [2 mg/ ml BSA, 0.9 % NaCl, protease inhibitor]. Ct25 and Ct28 test samples were prepared by dilution of the Ct21 test sample in the dilution buffer. Samples were aliquoted (120 µl), immediately frozen on dry ice and stored at - 80°C. For AgPOCT testing, samples were freshly thawed on ice before use. Test samples were validated using the SARS-CoV-2 Antigen Test by LumiraDx.

### AgPOCTs evaluated in this study

We included a total of 32 AgPOCTs available at local supermarkets, pharmacies and drugstores as well as on several online trade platforms (Table 1). Specific AgPOCTs will be referred to as the respective manufacturer’s name (in bold in Table 1). The inspected AgPOCTs include both, tests for professional *in vitro* diagnostics use (#1-14) as well as tests temporarily licensed for self-testing in Germany (#15-32) by the Federal Institute for Drugs and Medical Devices (Bundesinstitut für Arzneimittel und Medizinprodukte (BfArM); Supplemental Figure S5). The majority of AgPOCTs available were nasal or nasopharyngeal swab tests with the exception of BTNX, Ritter, Joinstar, Realy (#11-14) among the tests for professional use and Sanicom, Hygisun, fameditec (#30-32) among the self tests, which are all saliva spit tests, as well as Watmind (#29), which is a saliva swab test.

**Table 1:** AgPOCTs investigated in this study. For each AgPOCT supplier, name, reference and LOT number are indicated. If tests obtained a temporary license for self-testing in Germany the corresponding BfArM GZ number is given as well. In addition, sample type and professional (pro) versus layman (lay) use is indicated. In the last column the type of distributor where AgPOCTs were purchased is noted.

| # | Supplier | AgPOCT name | Specifications | Sample type | Use | Distributor |
| --- | --- | --- | --- | --- | --- | --- |
| 1 | **Abbott** Rapid Diagnostics Jena GmbH | Panbio COVID-19 Ag Rapid test device (nasal) | REF: 41FK11<br>LOT: 41ADG244A | Nasal swab | pro | Online trade |
| 2 | **Healgen** Scientific Limited Liability Company | Coronavirus Ag Rapid Test Cassette (Swab) | REF: GCCOV-502a<br>LOT: 2012650 | Nasopharyngeal swab | pro | Online trade |
| 3 | **RapiGEN**, INC. | Biocredit COVID-19 Ag – One step Rapid Test | REF: G61RHA20<br>LOT: H073097SD | Nasopharyngeal swab | pro | Online trade |
| 4 | Beijing **Beier** Bioengineering Co., Ltd | COVID-19 Antigen Rapid Test Kit | REF: not specified<br>LOT: 20210201 | Nasopharyngeal swab | pro | Online trade |
| 5 | **möLab** GmbH | mö-screen Testkit Corona Antigen | REF: 0230005B1<br>LOS: 2104072 | Nasal/<br>nasopharyngeal swab | pro | Online trade |
| 6 | **Biomerica**, Inc. | COVID-19 Antigen Rapid Test | REF: 1509A-25I<br>LOT: COV6686 | Nasopharyngeal swab | pro | Online trade |
| 7 | **Joysbio** (Tianjin) Biotechnology Co., Ltd | JOYSBIO SARS-CoV-2 Antigen Rapid Test Kit (Colloidal Gold) | REF: G10313<br>LOT: 2021011607 | Nasopharyngeal swab | pro | Online trade |
| 8 | **Safecare** Biotech (Hangzhou) Co., Ltd | COVID-19 Antigen Rapid Test Kit (Swab) | REF: COV Ag-6012<br>LOT: COV21040606 | Nasopharyngeal swab | pro | Online trade |
| 9 | Hangzhou **Testsea** Biotechnology Co., Ltd | Testsealabs COVID-19 Antigen Test Cassette | REF: 2020013 vB/10<br>LOT: TL1C05 | Nasal swab | pro | Online trade |
| 10 | **ACON** Biotech (Hangzhou) Co., Ltd | Flowflex SARS-Cov-2-Antigen-Schnelltest (Selbsttest) | REF: L031-11855<br>LOT: COV1030052 | Nasal swab | pro | Online trade |
| 11 | **BTNX** Inc. | Rapid Response COVID-19 Antigen Rapid Test Cassette | REF: COV-2C25B<br>LOT: COVG21030089 | Saliva (spit) | pro | Online trade |
| 12 | Joysbio (Tianjin) Biotechnology Co., Ltd/<br>**Ritter** | Easy Check Spit test SARS-CoV-2 Antigen Rapid Test Kit (Colloidal Gold) | REF: COV-AG-20/ G10313<br>LOT: 20210202 | Saliva (spit) | pro | Online trade |
| 13 | **Joinstar** Biomedical Technology Co., Ltd. | COVID-19 Antigen Rapid Test (Latex) | REF: RPBH12360<br>LOT: COV2103002L | Saliva/ sputum (spit), stool | pro | Online trade |
| 14 | Hangzhou **Realy** Tech Co., Ltd | Novel Coronavirus (SARS-CoV-2) Antigen Rapid Test Device (Saliva) | REF: K590516D<br>LOT: 202101022 | Saliva (spit) | pro | Online trade |
| 15 | **nal von minden** GmbH | NADAL COVID-19 Ag Test | REF: 243103N-20H<br>LOT: 175363<br>BfArM GZ: 5640-S-045/21 | Nasal swab | lay | Online trade |
| 16 | **SD Biosensor** | SARS-CoV-2 Rapid Antigen Test | REF: 9901-NCOV-01G<br>LOT: QCO390092I<br>BfArM GZ: 5640-S-025/21 | Nasal swab | lay | Online trade |
| 17 | Beijing **Hotgen** Biotech Co., Ltd | Coronavirus (2019-nCoV)-Antigentest | REF: 4260220532859<br>LOT: W2021032500/<br>W2021032602/ 1500<br>BfArM GZ: 5640-S-057/21 | Nasal swab | lay | Supermarket<br>Pharmacy |
| 18 | Guangzhou **Wondfo** Biotech Co., Ltd. | 2019-nCoV Antigen Test (Lateral Flow Method) | REF: W634P0021<br>LOT: W634104116<br>BfArM GZ: 5640-S-179/21 | Nasal swab | lay | Supermarket |
| 19 | **Teda** Laukoetter Technology GmbH | COVID-19 Antigen Schnelltest (kolloidales Gold) ANBIO Corona Antigen Nasentupfer | REF: A6061214<br>LOT: 2021046133/<br>461310/036138<br>BfArM GZ: 5640-S-079/21 | Nasal swab | lay | Drug store |
| 20 | Beijing **Lepu medical** Technology Co., Ltd | NASOCHECKcomofort SARS-CoV-2 Antigen-Schnelltest | REF: CG2701N<br>LOT: 21CG2720X/ 18X<br>BfArM GZ: 5640-S-104/21 | Nasal swab | lay | Drug store |
| 21 | Hangzhou **Clongene** Biotech Co., Ltd | COVID-19 Antigen Rapid Test | REF 6950921302636<br>LOT: 2021030161<br>BfArM GZ: 5640-S-168/21 | Nasal swab | lay | Online trade |
| 22 | Hangzhou **Laihe** Biotech Co., Ltd | LYHER Novel Coronavirus (COVID-19) Antigen Test Kit (Colloidal Gold) NASAL | REF: 303036<br>LOT: 2103049/47/ 89-01<br>BfArM GZ: 5640-S-009/21 | Nasal swab | lay | Pharmacy |
| 23 | **MP** Biomedicals Germany GmbH | Rapid SARS-Cov-2 Antigen Test Card | REF: 07AG6001BS<br>LOT: 21033003<br>BrArM GZ: 5640-S-076/21 | Nasal swab | lay | Supermarket |
| 24 | Xiamen **Boson** Biotech Co., Ltd | Rapid SARS-CoV-2 Antigen Test Card | REF: 1N40C5-4<br>LOT: 21040609<br>BfArM GZ: 5640-S-007/21 | Nasal swab | lay | Supermarket |
| 25 | **NanoRepro** AG | VIROMED for the detection of SARS-Cov-2 from anterior nasal swab | REF: B60500<br>LOT: 20210401B<br>BrArM GZ: 5640-S-096/21 | Nasal swab | lay | Drug store |
| 26 | Anhui **Deepblue** Medical Technology Co., Ltd | COVID-19 (SARS-CoV-2) Antigentestkit (kolloidales Gold) | REF: SL030101N-5<br>LOT: ST210405<br>BfArM GZ: 5640-S-086/21 | Nasal swab | lay | Online trade |
| 27 | **OFM** GmbH | Deni COVID-19 Antigen Test – Selbsttest für Zuhause | REF: OFM-LSYBT-NS-1<br>LOT: P202103003<br>BfArM GZ: 5640-S-140/21 | Nasal swab | lay | Drug store |
| 28 | **Medice** Arzneimittel Pütter GmbH & Co. KG | Medicovid-AG SARS-CoV-2 Antigen Selbsttest 5 NASE | REF: 1N40C5-4<br>LOT: 21041002<br>BfArM GZ: 5640-S-128/21 | Nasal swab | lay | Online trade |
| 29 | Shenzhen **Watmind** Medical Co., Ltd | SARS-CoV-2 Antigen Schnelltest zur Eigenanwendung (kolloidales Gold) | REF: LFA0401-5N<br>LOT: 21040904/ 21040704<br>BfArM GZ: 5640-032/21 | Saliva (swab) | lay | Supermarket |
| 30 | MR **Sanicom** GmbH | COVID-19 Antigen Schnelltest zur Eigenanwendung (Speichel-/ Spucktest) | Barcode no:<br>4260729310002<br>LOT: CAG2104021G<br>BfArM GZ: 5640-S-147/21 | Saliva (spit) | lay | Drug store |
| 31 | **Hygisun** Anbio<br>(Xiamen) Biotechnology<br>Co., Ltd. | COVID-19 Antigen Schnelltest<br>(kolloidales Gold) | REF: A6061213<br>LOT: 2021046132/<br>2021036136/ 2021036137<br>BfArM GZ: 5640-S-058/21 | Saliva (spit) | lay | Drug store |
| 32 | **fameditec** | CORA Check-19 Comfort | REF: K590516D/<br>LOT: 2021022019<br>BfArM GZ: 5640-S-154/21 | Saliva (collected<br>with sponge) | lay | Online trade |

For Lepu medical (#20 in Table 1), the AgPOCT with poorest results in our study, we purchased different versions and additional batches for a more in-depth characterization (Table 2).

**Table 2:** Lepu medical AgPOCT products investigated in this study. AgPOCT products made by Beijing Lepu Medical Technology Co., Ltd. are referred to as Lepu medical AgPOCTs. Lepu medical AgPOCTs were purchased on different online trade platforms. For each Lepu medical AgPOCT, product name, packaging size and intended use (professional (pro) versus layman (lay) use), reference/ barcode number, LOT number as well as BfArM GZ number if applicable are indicated. Production and use-by date are noted in the last column.

| Product name | Packaging size (use) | Specifications | Production and use-by date |
| --- | --- | --- | --- |
| SARS-CoV-2 Antigen Rapid Test Kit (Colloidal Gold Immunochromatography) CE | Pack of 25 tests/ (pro) | Barcode: 6921807601020<br>LOT: 21CG2713X | Production date: 06/03/2021<br>Use-by date: 06/03/2022 |
|  |  | Barcode: same as above<br>LOT: 21CG2715X | Production date: 03/11/2021<br>Use-by date: 03/11/2022 |
| NASOCHECKcomfort SARS-CoV-2 Antigen Schnelltest Immunchromatographischer Test (kolloidales Gold). Schnell & angenehm! Einfache Anwendung im vorderen Nasenbereich. | Pack of 25 testes (lay) | REF: CG2725(N)<br>Barcode: 4260716970042<br>LOT: 21CG2722X<br>BfArM GZ: 5640-S-104/21 | Production date: 31/03/202<br>Use-by date: 31/03/2022 |
| NASOCHECKcomfort SARS-CoV-2 Antigen-Schnelltest. Zur Eigenanwendung. Schnell & angenehm! Einfache Anwendung im vorderen Nasenbereich. | Single pack (narrow) (lay) | REF: CG2701N<br>Barcode: 4260716970059 LOT: 21CG2727X<br>BfArM GZ: 5640-S-104/21 | Production date: 11/04/2021<br>Use-by date: 11/04/2022 |
| NASOCHECKcomfort SARS-CoV-2 Antigen-Schnelltest. Zur Eigenanwendung. Schnell & angenehm! Einfache Anwendung im vorderen Nasenbereich. | Single pack (thick) (lay) | REF: CG2701N<br>Barcode: 4260716970059 LOT: 21CG2720X<br>BfArM GZ: 5640-S-104/21 | Production date: 26/03/2021<br>Use-by date: 26/03/2022 |
|  |  | REF, barcode, BfArM GZ: same as above<br>LOT: 21CG2724X | Production date: 04/04/2021<br>Use-by date: 04/04/2022 |

## Results

### Generation of test samples for standardized AgPOCT evaluation

In the present study, we sought to establish a standardized procedure to rapidly assess the sensitivities of a large number of SARS-COV-2 AgPOCTs. To this end, we generated a collection of test samples from pooled nasopharyngeal swabs from SARS-CoV-2 positive tested and negative tested individuals. Ct values of the SARS-CoV-2 positive pools were determined by RT-qPCR and test samples were prepared accordingly. The test sample collection comprised four SARS-CoV-2 positive pools with defined viral loads (Ct16, Ct21, Ct25, Ct28) and one SARS-CoV-2 negative pool. Per test sample, >200 aliquots with 120 µł sample volume each were prepared, allowing a quick and standardized evaluation of the analytical sensitivities of a large number of different AgPOCTs.

We estimated that our test sample collection covers a range from ∼7.0^*^10^8^ (Ct=16) to ∼1.7^*^10^5^ (Ct=28) SARS-CoV-2 genome copies per ml (Supplemental Table S6). We qualitatively validated our test sample collection using the LumiraDx SARS-CoV-2 Ag Test device, which was shown to have a high analytical sensitivity (Krüger *et al*., 2021). We used 50 µl of a test sample, each for the LumiraDx analysis and for all AgPOCTs evaluated in this study, as described before (Corman *et al*., 2021; Puyskens *et al*., 2021). All four SARS-CoV-2 positive test samples tested positive for SARS-CoV-2, while the negative test sample was recognized as negative in the LumiraDx analysis.

### Quantitative and qualitative assessment of AgPOCT analytical sensitivity

We tested a total of 32 AgPOCTs (Table 1). 12 AgPOCTs were purchased from local resellers (pharmacies, drugstores, supermarkets). Another 20 tests were purchased online. We performed the tests over 10 days, with the help of four students, during the course of four weeks. Freshly thawed aliquots of the Ct21, Ct25, and Ct28 test samples as well as the negative sample were used. We conducted two to four replicates per product, and acquired images of each of the tests at the time points specified by the manufacturers. The Ct16 test sample was only used for AgPOCTs that had low performance with the Ct21 test sample. For quantitative evaluation, signal intensities of the test (T) and the control (C) bands were measured and the ratio of these values (T/C ratio) was determined (Figure 1A). In addition, we scored a binary (positive or negative) test result using visual inspection of the images by four different persons (Figure 1B).

**Figure 1:**
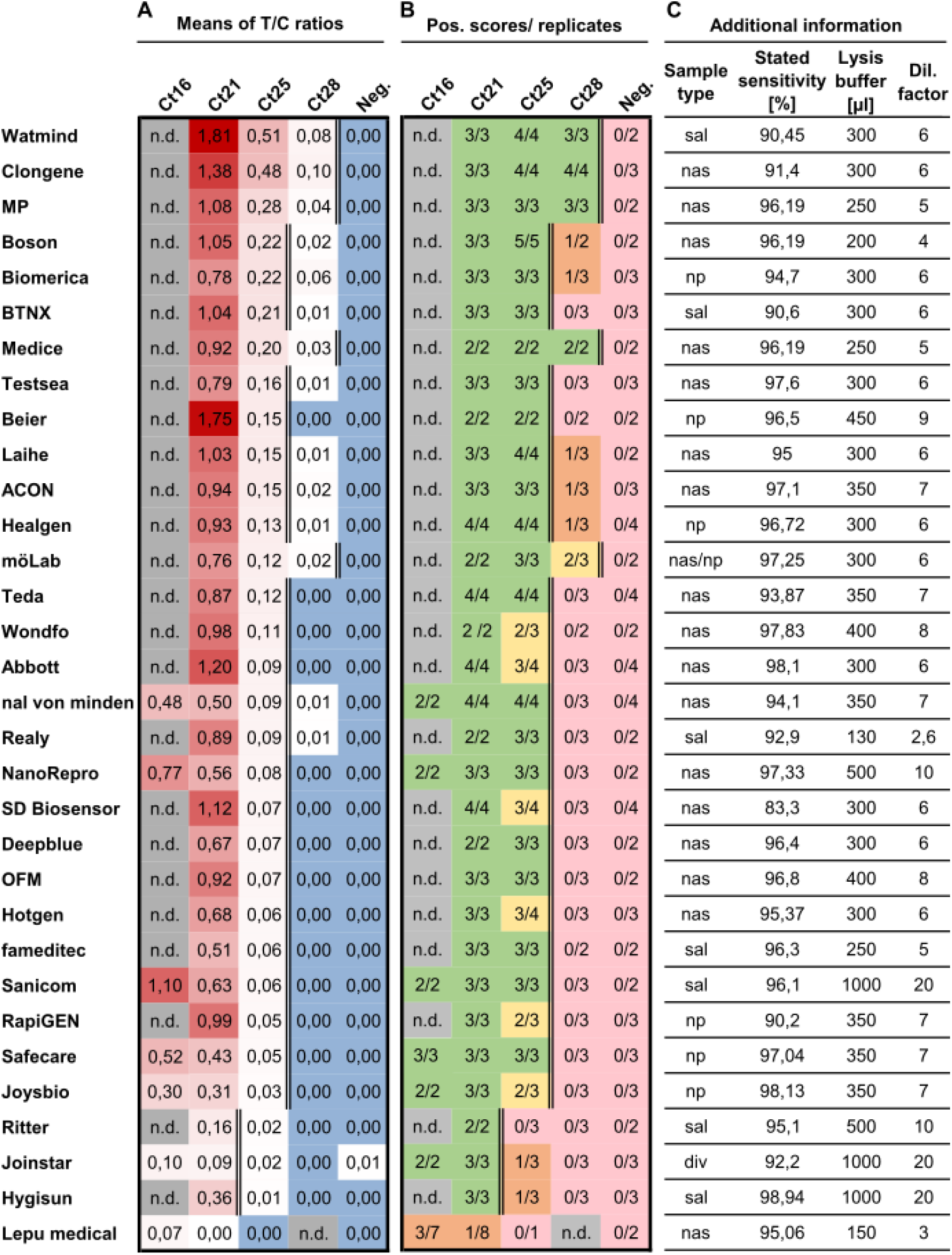
Quantitative and qualitative evaluation of 32 SARS-CoV-2 AgPOCTs in a rapid sensitivity assessment approach. (A) Investigated AgPOCT are listed with the means of T/C ratios (test band (T) intensity to control (C) band intensity) for each Ct test sample. T/C ratios are color-coded in shades of red (highest values with most intense red). Blue color highlights zeros indicating the absence of measurable signal at the test band position. Ct16 test sample was only used on AgPOCTs with exceptionally low performance in detection of the Ct21 sample. AgPOCTs are ranked according to their T/C_Ct25_ ratio. (B) Scoring results of visual inspection for all replicates. Full reproducibility of positive scores in all replicates is highlighted in green, positive scores in the majority of replicates in yellow, positive scores in the minority of replicates in orange and no positive scores in any replicate in light red. n.d. = not determined (grey). Double line indicates the limit of reliable detection of SARS-CoV-2 positive samples (reliability defined by reproducibility of positive scores in all (green) or most (yellow) replicates of a given Ct test sample). (C) Additional information on investigated AgPOCTs: Sample type (nasal (nas)/ nasopharyngeal (np) swab, saliva (sal) or diverse (div)), sensitivities of AgPOCTs according to the corresponding manufacturer’s package insert, volumes of provided lysis buffer and the resulting dilution factor for the Ct test samples (V = 50 µl) are given.

**Figure 2:**
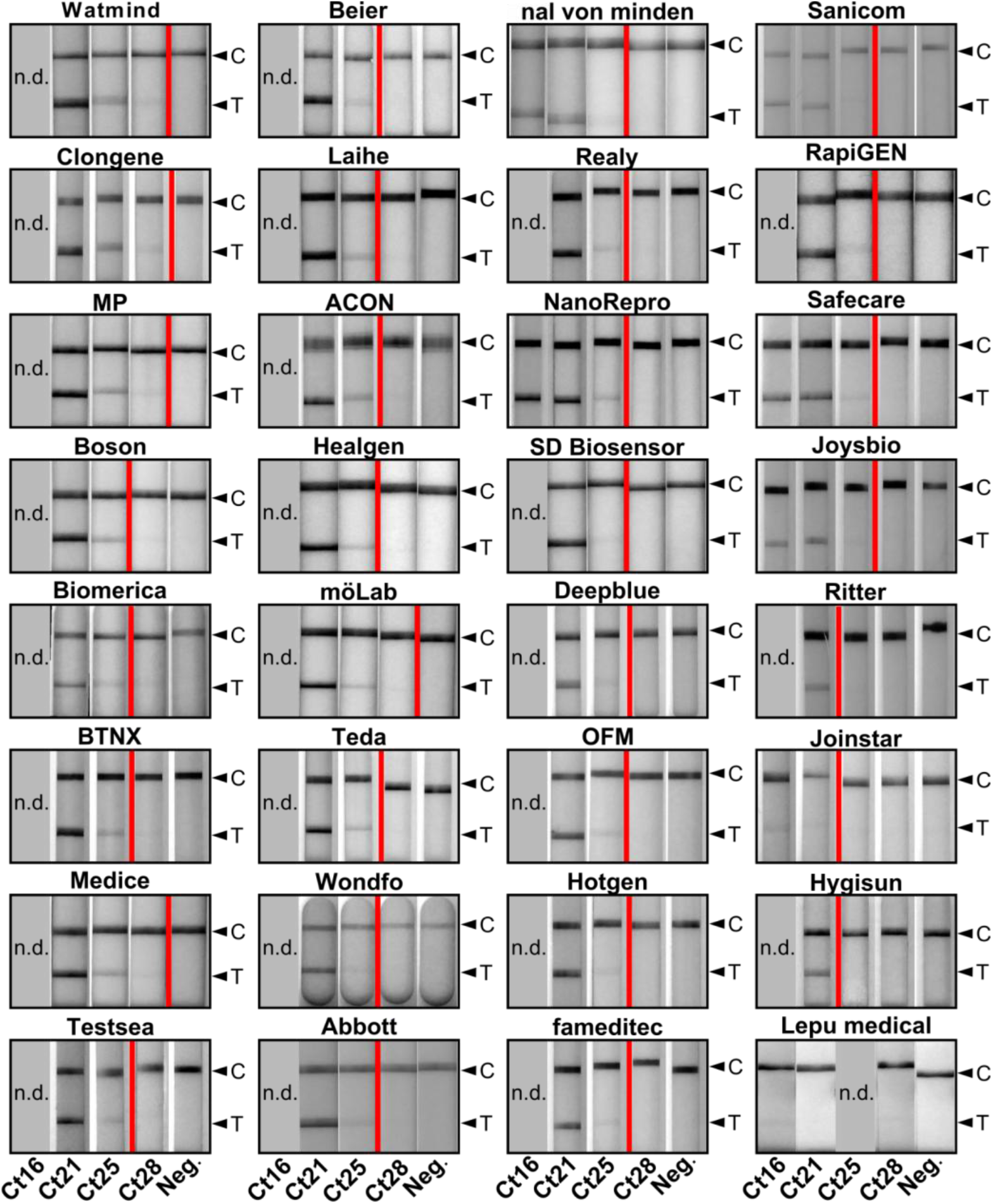
Representative images of SARS-CoV-2 AgPOCTs lateral flow test stripes treated with corresponding Ct test samples. Contrast settings were optimized for each AgPOCT example image set in order to ensure best visibility of the test bands. AgPOCT example images are arranged (from top left to bottom right) according to the ranking presented in Figure 1. Red line indicates the limit of reliable detection (see Figure 1A, B). Arrowheads highlight positions of control (C) and test (T) band.

For 31 of 32 investigated AgPOCTs, an average T/C_Ct25_>0 was determined for all virus-containing samples and not for the negative control sample (Figure 1A). This indicates that the digital quantification detects test band signals for 31 AgPOCTs using the Ct25 test sample, albeit sometimes with extremely weak signal intensities. Only for Jointstar, one replicate of the negative test sample resulted in a false positive test band indicated by a T/C_Neg._>0. In contrast to the more sensitive digital quantification, visual inspection did only score a positive result for 28 of 31 AgPOCTs with a T/C_Ct25_>0 (Figure 1B). This also holds true for the visual assessment of the results of technical replicates, e.g. for Jointstar, the negative sample with a T/C_Neg._>0 scored negative in the visual inspection. We could not establish a specific T/C value threshold to explain the results of the visual assessment, indicating that these ratios are product-specific. This can be explained by different dyes and dye-systems, and by the fact that the visual assessment was conducted using color vision, while for T/C quantification grayscale images were used. We also observed a large coefficient of variation (CV) for some of the tests, in particular for samples with very small T/C ratios, emphasizing weak signals close to the detection limit of the digital quantification (Supplemental Figure S1C).

We grouped the tested AgPOCTs into categories with low (Group III), medium (Group II) and high (Group I) sensitivity based on the reliability to detect a given SARS-CoV-2 positive sample. A sample was considered reliably detected by a given AgPOCT when all or the majority of replicates (at least two out of three or three out of four replicates) of a given sample were scored positive. If none or the minority of replicates of a given sample was detected by the corresponding AgPOCT, reliability requirements were not met.

One exception was Lepu medical (Table 1, AgPOCT #20; Table 2, last row) which did not fulfill the requirements for any of these groups. For Group III AgPOCTs with the lowest sensitivity, the minimum criterion was that the tests were able to reliably detect the Ct21 sample, and the Lepu medical tests failed this, as they did not even reliably score positive with the Ct16 sample (∼7.0^*^10^8^ copies/ ml; Figure 1B). To investigate this product further we used individual unprocessed nasal/ nasopharyngeal swab samples with low Ct values (Ct13.3 to Ct18.4) on this and another poor performing product. Comparison of these results to the Ct16 test sample confirmed the low sensitivity of the Lepu medical (Supplemental Figure S2). Only for samples with very low Ct values (Ct∼13) T/C ratios were obtained that can be detected easily visually (Supplemental Figure S2A). This suggests that this product is not completely non-functional, but largely insensitive. Since Lepu medical AgPOCTs have been widely used in Germany and other European countries we retrieved several Lepu medical products available on different online trade platforms (Table 2). These included two different batches of a Lepu medical product intended for professional use (Table 2, first row, no BfArM GZ number, CE mark). Additionally, we included different batches and deviations of the Lepu medical AgPOCT described before (Table 1, AgPOCT #20; Table 2, last three rows). These products were provided with the same BfArM GZ number (5640-S-104/21), which cannot be found on the BfArM list anymore (Supplemental Figure S5). We investigated their performance in direct comparison in multiple replicates using Ct16, Ct21 and Ct25 test samples (Supplemental Figure S3). This revealed high variation of the determined T/C ratios, with coefficients of variation (CV) ranging from 0.26 to 1.54 and a median CV of 0.50 (Supplemental Figure S4C). In contrast, the median CV of the other 31 investigated AgPOCTs with Ct16-25 test samples was 0.11 (Supplemental Figure S1C). This indicates a larger variability of the test results not only for different implementations of the Lepu medical AgPOCTs, but also for different batches of the same Lepu medical product, compared to all other AgPOCTs investigated in this study.

AgPOCTs in Group III only reliably detected the Ct21 sample (∼2.2^*^10^7^ copies/ ml) and include Hygisun, Joinstar and Ritter. Of note is that all of them are saliva based spit tests (Table 1), which are provided with a considerably larger amount of lysis buffer (500-1000 µl lysis buffer; Figure 1C) than most other AgPOCTs resulting in an increased dilution of the test sample compared to AgPOCTs for nasal samples, which are provided on average with 320 µl lysis buffer (Figure 1C). The resulting higher dilution of the sample together with the possibility of lower virus concentration in saliva versus nasal or nasopharyngeal swabs may further influence the sensitivity of these saliva tests.

The large majority of the investigated AgPOCTs (23 out of 32) delivered visible positive results with the Ct25 sample (∼1.4^*^10^6^ copies/ ml, Group II). Among these 23 AgPOCTs, positive scoring was fully reproducible in all replicates for 17 AgPOCTs. AgPOCTs intended for professional use (sorted ascending according to T/C_Ct25_: Safecare, Realy, Healgen, ACON, Beier, Testsea, BTNX and Biomerica) largely cluster in the upper half of the T/C_Ct25_ ranking, while tests licensed for self-testing largely cluster in the lower half (sorted ascending according to T/C_Ct25_: Sanicom, fameditec, OFM, Deepblue, NanoRepro, nal von minden, Teda, Laihe and Boson). Interestingly, among both tests for professional and for layman use, saliva spit tests (Realy, Sanicom, fameditec) appear largely inferior compared to nasal swab tests in this setting with the exception of BTNX, which is the sixth highest ranked AgPOCT among all investigated tests. Six AgPOCTs in Group II (sorted ascending according to T/C_Ct25_: Joysbio, RapiGEN, Hotgen, SD Biosensor, Abbott and Wondfo) failed in one out of three to four replicates to detect the Ct25 sample, which is represented by larger CV values ranging from 0.26 to 0.87 (Supplemental Figure S1C). Using the Ct28 test sample (∼1.7^*^10^5^ copies/ ml), 14 out of 32 AgPOCTs yielded a T/C_Ct28_>0, however, only five reliably scored positive in the visual investigation (Group I). These include (sorted in ascending order according to T/C_Ct25_) möLab, Medice, MP, Clongene and Watmind, three of which are temporarily licensed for self-testing (Table 1, Supplemental Figure S5). All, except möLab delivered a positive visual result in all three replicates.

Taken together, the data presented here demonstrate that the different SARS-CoV-2 AgPOCTs available deviate largely in the analytical sensitivity of the lateral flow test stripes and provided buffer systems, corresponding more than two orders of magnitude of viral genome copies per ml (7.0^*^10^8^ to 1.7^*^10^5^). Additionally, we revealed a great variability in results delivered with different Lepu medical AgPOCT versions and batches emphasizing the need for regular quality monitoring.

## Discussion

We developed a straight-forward strategy to evaluate the technical sensitivity of AgPOCTs for SARS-CoV-2. Using a set of four SARS-CoV-2 positive reference samples spanning the relevant dynamic range of the typical sensitivity of AgPOCTs (∼1.7^*^10^5^ to ∼2.2^*^10^7^ SARS-CoV-2 genome copies per ml) we were able to group 32 commercially available products into AgPOCT groups with high, average and low sensitivity (Group I-III). Most importantly, we identified one product that did not detect any of the test samples and therefore is considered not suitable for SARS-CoV-2 diagnostics.

The majority of tests investigated in this study reliably detected the Ct25 test sample as SARS-CoV-2 positive (Group II). Some of these AgPOCTs have been thoroughly characterized, including Abbott, RapiGEN, Healgen, nal von minden and SD Biosensor by Corman and colleagues (Corman *et al*., 2021) among others (Strömer *et al*., 2021; Stokes *et al*., 2021; Merino *et al*., 2021; Schildgen *et al*., 2021; Seynaeve *et al*., 2021; Nordgren *et al*.; Puyskens *et al*., 2021; Scheiblauer *et al*., 2021; Kohmer *et al*., 2021; Wagenhäuser *et al*., 2021; Berger *et al*., 2021; Jegerlehner *et al*., 2021; Iglòi *et al*., 2021; Bekliz *et al*., 2021; Cubas-Atienzar *et al*., 2021, Haage *et al*., 2021 and more). Corman and colleagues determined 95% limits of detection for each AgPOCT using 138 SARS-CoV-2 positive clinical samples with viral loads ranging from 1.9^*^10^4^ to 2.8^*^10^9^ genome copies per ml. Among the AgPOCTs also tested in this study, Healgen was found to be most sensitive closely followed by Abbott, SD Biosensor and nal von minden - all with a 95% limit of detection between 2.3 - 9.3^*^10^6^ SARS-COV-2 genomes per swab. In contrast, for RapiGEN a 95% limit of detection more than three orders of magnitudes lower was found. This discrepancy in performance between RapiGEN and the above mentioned products is supported by other studies (Brümmer *et al*., 2021). In our analysis, this trend is also reflected even though we cannot resolve the limits of detection in such great detail: For Healgen and nal von minden, detection of the Ct25 test sample (∼1.4^*^10^6^ copies/ ml) was robust with all replicates being positively scored. For RapiGEN, Ct25 test sample detection was less reliable and based on the T/C_Ct25_, this product is ranked in the lowest quarter among all AgPOCTs investigated.

Among the 32 investigated AgPOCTs, we identified four reliably well performing AgPOCTs, which detected the Ct28 test sample (∼1.7^*^10^5^ copies/ ml) as SARS-CoV-2 positive in all replicates (Group I). These include in ascending order (based on T/C_Ct25_) Medice, MP, Clongene and Watmind. The latter represents the test winner in our study and is also among the best three AgPOCTs out of 122 tested products with a sensitivity of 82 % in samples with Ct values ranging from 17 to 35 corresponding to viral loads of >10^8^ to 10^3^ SARS-CoV-2 genome copies per ml (Scheiblauer *et al*., 2021).

Group III includes AgPOCTs with low performance as these only detected the Ct21 test sample as SARS-CoV-2 positive. For Joinstar, using Latex beads for visualisation, evidence provided by Scheiblauer and colleagues (2021) suggests that this test is non-functional with 0% sensitivity for all sample panels supporting the low ranking of Joinstar in this study. In our analysis we detected very weak bands for the Ct21 and Ct16 test samples, however, these were considerably weaker than for all other tests suggesting the possibility that latex beads used for visualisation do fail to produce a strong signal. Besides Joinstar, Ritter and Hygisun, both saliva spit tests similar to Joinstar, also showed low sensitivity in our studies. While we could not find independent evaluation studies for these products, both can be found on the BfArM list (as of July 23, 2021; Supplemental Figure S5).

Among the low ranked AgPOCTs, the sensitivity of Lepu medical AgPOCT was exceptionally low as this test failed to deliver a visible positive test result in most replicates, even for the Ct16 sample. In addition to its poor performance in SARS-CoV-2 diagnostics, out of 20 performed Lepu medical tests three tests technically failed, indicated by the absence of the control band. Importantly, this last test is a very popular product in the area where this study was conducted, available at many drugstores and supermarket chains. Of note is also that Lepu medical differs from other AgPOCTs in its design and sample application. Technical failure did not occur in any of the other AgPOCTs, in which the immunochromatography paper is embedded in the common plastic cassettes. In other studies tested Lepu medical AgPOCT products performed better (Scheiblauer *et al*., 2021; Baro *et al*., 2021), e.g. in the setting of Scheiblauer *et al*. (2021), a sensitivity of 100 % was found for a Lepu medical AgPOCT and test panel members with Ct values ranging from Ct17 to Ct25. As the AgPOCTs used in these studies are not specified with reference/product and LOT number, it is possible that a different Lepu medical product or batch was used. We purchased different Lepu medical AgPOCT products available online and compared performances on the pooled test samples as well as on raw, unprocessed swab samples (Supplemental Figure 3). We tested two batches of a CE-marked product and four Lepu medical with the same BfArM GZ number, but different packaging sizes (Table 2). Indeed, we identified two Lepu medical products performing better than shown in Figure 1, however, these performances were not reproducible with other batches of the same product (Supplemental Figure 3) indicating batch-specific variation of the quality. This again emphasizes the importance of a simple method to assay the performance of a product and corresponding batches. Taken together, comparison with published data for some of the investigated products confirmed our results. Therefore, we provide evidence that our chosen strategy constitutes a viable solution to rapidly assess the sensitivity of SARS-CoV-2 AgPOCTs.

It is important to mention that the sensitivity of an AgPOCT is the product of multiple factors; the sensitivity of the test stripe and buffer system used are important contributing factors, but not the only ones. Another is the volume of the lysis buffer provided with each AgPOCT. The volume varies between tests of different manufacturers resulting in a 2.6-to 20-fold dilution of the test samples during the procedure (Figure 1C). In this study, test results were not corrected for these different dilution factors, because the sample dilution is an internal property of each AgPOCT. By using the same sample volume for each AgPOCT we also neglect potential differences in swab properties, such as absorption volume or sample specimen (saliva, nasal or nasopharyngeal samples), which affect the diagnostic sensitivity of AgPOCTs. However, we note that for tests based on nasal swabs the used volume of 50 μl approximates the quantity absorbed by these swabs (Corman *et al*., 2021). Furthermore, the AgPOCT-specific instructions for self-sampling, which will influence how careful a sample is collected, can also influence the diagnostic sensitivity of a test. In light of these considerations we want to emphasize that this evaluation method only and exclusively focuses on comparing the technical sensitivity of the lateral flow test strips from different test manufacturers, in combination with the provided lysis buffers.

Currently, there are more than 500 different products available for SARS-CoV-2 diagnostics, many of which lack independent assessment of their performance. In most cases the clinical sensitivity values provided by the manufacturer (e.g. in Figure 1C) are far >90% (Figure 1C). However, detailed information on specimen collection and viral loads are usually not provided rendering these values largely inconclusive and misleading for laymen. Considering that individual products use different antibodies in varying amounts with different specificities and affinities sometimes recognizing different proteins in the viral particle with differing abundances, and diverse staining methods, these conspicuously similar values for clinical sensitivity given by the manufacturer are also unlikely. Therefore, an independent, rapid and critical evaluation of AgPOCTs available is required in order to determine the realistic performance of AgPOCT relevant to the daily user and especially to identify poor performing products. Given the huge number of products available for rapid SARS-CoV-2 diagnostics, in-depth studies evaluating the quality of AgPOCTs in a time-intensive procedure will not be available any time soon for all products available. Therefore, the procedure presented here involving a reduced test sample collection and minimal labor represents a feasible strategy for prompt evaluation of available AgPOCTs for their usability in SARS-CoV-2 diagnostics. We provide a useful estimation of the limits of detection for the investigated AgPOCTs as the dimensions and trends are comparable to results from much more laborious in-depth studies. Importantly, using this approach we revealed very heterogeneous results for the Lepu medical AgPOCT, which precludes in our opinion the use of this product (or product family) for SARS-CoV-2 diagnostics. In conclusion, we suggest this procedure as a rapid alternative to investigate Covid-19 AgPOCs in the absence of reliable data that validate the performance of a specific product and related batches.

## Supporting information

upplemental Figure S3

## Data Availability

All data referred to the manuscript is available upon request.

## Acknowledgements

We thank Helena Kettern, Vincent T. Jaschinski and Larissa Karl for their flexible help with AgPOCT image acquisition. Furthermore, we thank M. Krogemann for help with AgPOCT result scoring. Additionally, we thank M. Meurer for helpful

