## Supplemental Figure S1 for "Rapid comparative evaluation of SARS-CoV-2 rapid point-of-care antigen tests"

| | A Means of T/C ratios | | | | | B SD of T/C ratios | | | | | C CV of T/C ratios | | | | |
|---|---|---|---|---|---|---|---|---|---|---|---|---|---|---|---|
| | Ct16 | Ct21 | Ct25 | Ct28 | Neg. | Ct16 | Ct21 | Ct25 | Ct28 | Neg. | Ct16 | Ct21 | Ct25 | Ct28 | Neg. |
| Watmind | n.d. | 1,81 | 0,51 | 0,08 | 0,00 | n.d. | 0,10 | 0,05 | 0,02 | 0,00 | n.d. | 0,06 | 0,10 | 0,20 | n.d. |
| Clongene | n.d. | 1,38 | 0,48 | 0,10 | 0,00 | n.d. | 0,05 | 0,08 | 0,03 | 0,00 | n.d. | 0,04 | 0,16 | 0,32 | n.d. |
| MP | n.d. | 1,08 | 0,28 | 0,04 | 0,00 | n.d. | 0,09 | 0,04 | 0,01 | 0,00 | n.d. | 0,08 | 0,16 | 0,28 | n.d. |
| Boson | n.d. | 1,05 | 0,22 | 0,02 | 0,00 | n.d. | 0,00 | 0,04 | 0,00 | 0,00 | n.d. | 0,00 | 0,18 | 0,21 | n.d. |
| Biomerica | n.d. | 0,78 | 0,22 | 0,06 | 0,00 | n.d. | 0,24 | 0,01 | 0,03 | 0,00 | n.d. | 0,31 | 0,06 | 0,47 | n.d. |
| BTNX | n.d. | 1,04 | 0,21 | 0,01 | 0,00 | n.d. | 0,07 | 0,06 | 0,02 | 0,00 | n.d. | 0,07 | 0,29 | 1,73 | n.d. |
| Medice | n.d. | 0,92 | 0,20 | 0,03 | 0,00 | n.d. | 0,02 | 0,01 | 0,00 | 0,00 | n.d. | 0,02 | 0,06 | 0,17 | n.d. |
| Testsea | n.d. | 0,79 | 0,16 | 0,01 | 0,00 | n.d. | 0,24 | 0,02 | 0,02 | 0,00 | n.d. | 0,31 | 0,15 | 1,73 | n.d. |
| Beier | n.d. | 1,75 | 0,15 | 0,00 | 0,00 | n.d. | 0,05 | 0,04 | 0,00 | 0,00 | n.d. | 0,03 | 0,23 | n.d. | n.d. |
| Laihe | n.d. | 1,03 | 0,15 | 0,01 | 0,00 | n.d. | 0,06 | 0,01 | 0,01 | 0,00 | n.d. | 0,06 | 0,09 | 0,88 | n.d. |
| ACON | n.d. | 0,94 | 0,15 | 0,02 | 0,00 | n.d. | 0,18 | 0,01 | 0,01 | 0,00 | n.d. | 0,19 | 0,04 | 0,50 | n.d. |
| Healgen | n.d. | 0,93 | 0,13 | 0,01 | 0,00 | n.d. | 0,09 | 0,05 | 0,01 | 0,00 | n.d. | 0,09 | 0,39 | 1,73 | n.d. |
| möLab | n.d. | 0,76 | 0,12 | 0,02 | 0,00 | n.d. | 0,07 | 0,01 | 0,00 | 0,00 | n.d. | 0,09 | 0,09 | 0,13 | n.d. |
| Teda | n.d. | 0,87 | 0,12 | 0,00 | 0,00 | n.d. | 0,04 | 0,02 | 0,00 | 0,00 | n.d. | 0,05 | 0,18 | n.d. | n.d. |
| Wondfo | n.d. | 0,98 | 0,11 | 0,00 | 0,00 | n.d. | 0,05 | 0,09 | 0,00 | 0,00 | n.d. | 0,05 | 0,87 | n.d. | n.d. |
| Abbott | n.d. | 1,20 | 0,09 | 0,00 | 0,00 | n.d. | 0,10 | 0,07 | 0,00 | 0,00 | n.d. | 0,08 | 0,73 | n.d. | n.d. |
| nal von minden | 0,48 | 0,50 | 0,09 | 0,01 | 0,00 | 0,02 | 0,08 | 0,01 | 0,02 | 0,00 | 0,04 | 0,16 | 0,13 | 1,73 | n.d. |
| Realy | n.d. | 0,89 | 0,09 | 0,01 | 0,00 | n.d. | 0,05 | 0,01 | 0,01 | 0,00 | n.d. | 0,06 | 0,12 | 1,73 | n.d. |
| NanoRepro | 0,77 | 0,56 | 0,08 | 0,00 | 0,00 | 0,02 | 0,06 | 0,01 | 0,00 | 0,00 | 0,03 | 0,10 | 0,15 | n.d. | n.d. |
| SD Biosensor | n.d. | 1,12 | 0,07 | 0,00 | 0,00 | n.d. | 0,12 | 0,03 | 0,00 | 0,00 | n.d. | 0,11 | 0,35 | n.d. | n.d. |
| Deepblue | n.d. | 0,67 | 0,07 | 0,00 | 0,00 | n.d. | 0,01 | 0,00 | 0,00 | 0,00 | n.d. | 0,01 | 0,04 | n.d. | n.d. |
| OFM | n.d. | 0,92 | 0,07 | 0,00 | 0,00 | n.d. | 0,08 | 0,04 | 0,00 | 0,00 | n.d. | 0,09 | 0,53 | n.d. | n.d. |
| Hotgen | n.d. | 0,68 | 0,06 | 0,00 | 0,00 | n.d. | 0,18 | 0,02 | 0,00 | 0,00 | n.d. | 0,26 | 0,26 | n.d. | n.d. |
| fameditec | n.d. | 0,51 | 0,06 | 0,00 | 0,00 | n.d. | 0,05 | 0,02 | 0,00 | 0,00 | n.d. | 0,10 | 0,26 | n.d. | n.d. |
| Sanicom | 1,10 | 0,63 | 0,06 | 0,00 | 0,00 | 0,00 | 0,03 | 0,01 | 0,00 | 0,00 | 0,00 | 0,05 | 0,22 | n.d. | n.d. |
| RapiGEN | n.d. | 0,99 | 0,05 | 0,00 | 0,00 | n.d. | 0,05 | 0,05 | 0,00 | 0,00 | n.d. | 0,05 | 0,87 | n.d. | n.d. |
| Safecare | 0,52 | 0,43 | 0,05 | 0,00 | 0,00 | 0,05 | 0,03 | 0,01 | 0,00 | 0,00 | 0,11 | 0,07 | 0,22 | n.d. | n.d. |
| Joysbio | 0,30 | 0,31 | 0,03 | 0,00 | 0,00 | 0,04 | 0,04 | 0,01 | 0,00 | 0,00 | 0,13 | 0,12 | 0,36 | n.d. | n.d. |
| Ritter | n.d. | 0,16 | 0,02 | 0,00 | 0,00 | n.d. | 0,05 | 0,00 | 0,00 | 0,00 | n.d. | 0,30 | 0,17 | n.d. | n.d. |
| Joinstar | 0,10 | 0,09 | 0,02 | 0,00 | 0,01 | 0,05 | 0,03 | 0,03 | 0,00 | 0,02 | 0,49 | 0,30 | 1,73 | n.d. | 1,73 |
| Hygisun | n.d. | 0,36 | 0,01 | 0,00 | 0,00 | n.d. | 0,02 | 0,01 | 0,00 | 0,00 | n.d. | 0,07 | 0,88 | n.d. | n.d. |
| Lepu medical | 0,07 | 0,00 | 0,00 | n.d. | 0,00 | 0,06 | 0,01 | n.d. | n.d. | 0,00 | 0,90 | 2,83 | n.d. | n.d. | n.d. |

**Supplemental Figure S1: Color-coded representation of the standard deviation (SD) and coefficient of variation (CV) calculated for the T/C ratios of different AgPOCTs treated with different Ct test samples.** (A) Means of T/C ratios (test band (T) intensity to control (C) band intensity) for each AgPOCT and Ct test sample (reproduced from Figure 1). Double line indicates the limit of reliable detection of SARS-CoV-2 positive samples (see Figure 1A, B). (B) Standard deviations (SD) of T/C ratios for each AgPOCT and Ct test sample . (C) Coefficients of variation (CV) of T/C ratios for each AgPOCT and Ct test sample. Means, SD and CV are color-coded in shades of red (highest values with most intense color). Blue color highlights zeros. n.d. (grey) = not determined.
