## Supplemental Figure S2 for "Rapid comparative evaluation of SARS-CoV-2 rapid point-of-care antigen tests"

**A**

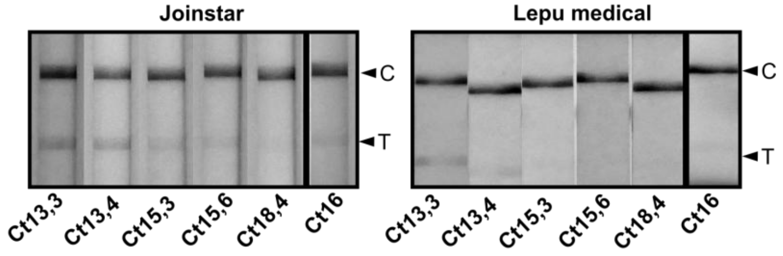


**B**

| | Unprocessed swabs samples: T/C ratios | | | | | Test sample: T/C ratios | Test sample: SD | Test sample: CV |
|---|---|---|---|---|---|---|---|---|
| | Ct13,3 | Ct13,4 | Ct15,3 | Ct15,6 | Ct18,4 | Ct16 | Ct16 | Ct16 |
| Lepu medical | 0,19 | 0,10 | 0,04 | 0,00 | 0,00 | 0,07 | 0,06 | 0,90 |
| Joinstar | 0,24 | 0,30 | 0,13 | 0,13 | 0,06 | 0,10 | 0,05 | 0,49 |

**Supplemental Figure S2: Testing of individual, unprocessed nasopharyngeal swab samples on low performing AgPOCTs.** (A) Representative images of lateral flow test stripes of Lepu medical and Joinstar with unprocessed and Ct16 test samples. Arrowheads highlight positions of control (C) and test (T) band. Thick black line separates unprocessed samples from Ct16 test sample. (B) Means of T/C ratios (test band intensity to control band intensity) for Lepu medical and Joinstar with unprocessed swab samples and Ct16 test sample (data for Ct16 test sample reproduced from Figure 1A). Standard deviations (SD) and coefficients of variation (CV) of T/C ratios are given for the Ct16 test sample. Means, SD and CV are color-coded in shades of red (highest values with most intense color). Blue color highlights zeros. n.d. = not determined (grey).
