## Supplementary material for "Rapid comparative evaluation of SARS-CoV-2 rapid point-of-care antigen tests": upplemental Figure S3

| Lepu medical test name | Reference no. LOT no. | A: Means of T/C ratios Ct16 | Ct21 | Ct25 | n | B: T/C ratio Ct21 (swab dip) | C: T/C ratios raw swab samples Ct13,3 | Ct13,4 | Ct15,3 | Ct16,0 |
|---|---|---|---|---|---|---|---|---|---|---|
| SARS-CoV-2 Antigen Rapid Test Kit (Colloidal Gold Immunochromatography) CE (pack of 25 tests) | 6921807601020 21CG2713X | 0,06 | 0,01 | 0,00 | 4-5 | 0,08 | 0,21 | 0,16 | 0,08 | 0,05 |
| | 6921807601020 21CG2715X | 0,12 | 0,06 | 0,00 | 4 | n.d. | 0,69 | 0,63 | 0,30 | 0,11 |
| NASOCHECKcomfort SARS-CoV-2 Antigen Schnelltest Immunchromatographischer Test (kolloidales Gold) (pack of 25 tests) | CG2725(N) 21CG2722X | 0,06 | 0,04 | 0,00 | 2-3 | 0,05 | 0,19 | 0,23 | 0,06 | 0,15 |
| NASOCHECKcomfort SARS-CoV-2 Antigen Schnelltest Zur Eigenanwendung (single pack; narrow) | CG2701N 21CG2727X | 0,10 | 0,04 | 0,00 | 2-3 | 0,05 | 0,34 | 0,16 | 0,03 | 0,03 |
| NASOCHECKcomfort SARS-CoV-2 Antigen Schnelltest Zur Eigenanwendung (single pack; thick) | CG2701N 21CG2720X | 0,08 | 0,04 | 0,00 | 4-5 | 0,04 | 0,11 | 0,12 | 0,02 | 0,00 |
| | CG2701N 21CG2724X | 0,13 | 0,04 | 0,00 | 4 | n.d. | 0,61 | 0,55 | 0,12 | 0,15 |

**D**

SARS-CoV-2 Antigen Rapid Test Kit (Colloidal Gold Immunochromatography) CE (pack of 25 tests) — 6921807601020 21CG2713X; 6921807601020 21CG2715X

NASOCHECKcomfort SARS-CoV-2 Antigen Schnelltest Immunchromatographischer Test (kolloidales Gold) (pack of 25 tests) — CG2725(N) 21CG2722X

NASOCHECKcomfort SARS-CoV-2 Antigen Schnelltest Zur Eigenanwendung (single pack; narrow) — CG2701N 21CG2727X

NASOCHECKcomfort SARS-CoV-2 Antigen Schnelltest Zur Eigenanwendung (single pack; thick) — CG2701N 21CG2720X; CG2701N 21CG2724X

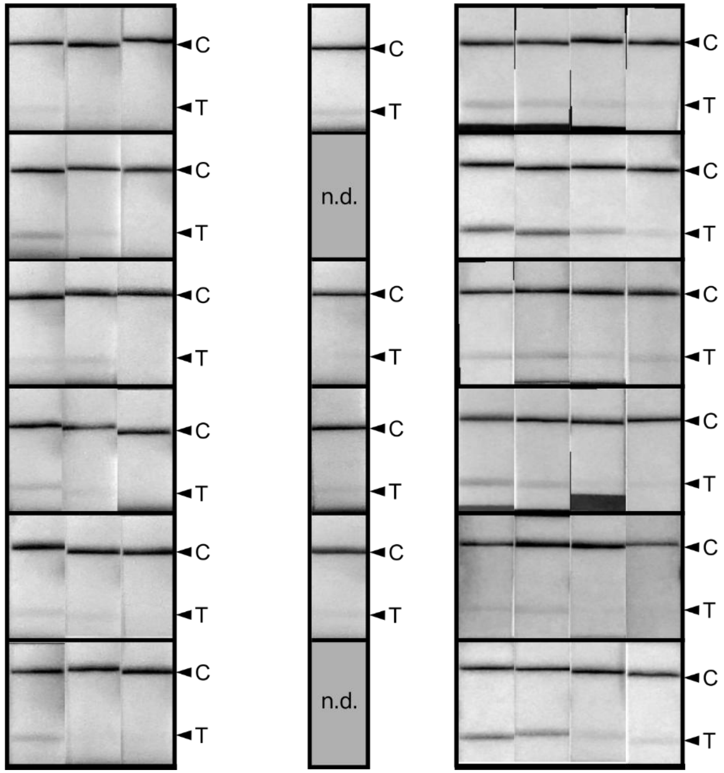


**Supplemental Figure S3: Comparison of different Lepu medical AgPOCT products.** *(continued on next page)*

**Supplemental Figure S3: Comparison of different Lepu medical AgPOCT products.** *(continued)* Different Lepu medical AgPOCT products are listed with reference/ barcode number and LOT number in the first column. The first two products did not have a BfArM GZ number and were CE-marked. Remaining products were provided with the BfArM GZ 5640-S-104/21. (A) Means of T/C ratios (test band (T) intensity to control (C) band intensity) for each Lepu medical AgPOCT product and Ct test sample (Ct16, Ct21, Ct25). Test samples were applied on swabs by pipetting. Numbers of replicates (n) are provided. (B) T/C ratios obtained for each Lepu medical AgPOCT and Ct21 test sample when swabs were dipped into the sample (n=1). (C) T/C ratios for each Lepu medical AgPOCT and unprocessed, raw swab samples in VTM with Ct values ranging from Ct13,3 to Ct16,0 (n=1). Values are color-coded in shades of red (highest values with most intense color). Blue color highlights zeros. n.d. (grey) = not determined. (D) Representative images of lateral flow test stripes of different Lepu medical products and corresponding samples (A-C). Arrowheads highlight positions of control (C) and test (T) band.
