## Supplemental Figure S4 for "Rapid comparative evaluation of SARS-CoV-2 rapid point-of-care antigen tests"

| Lepu medical test name | Reference no. LOT no. | A: Means of T/C ratios Ct16 | Ct21 | Ct25 | n | B: SD of T/C ratios Ct16 | Ct21 | Ct25 | C: CV of T/C ratios Ct16 | Ct21 | Ct25 |
|---|---|---|---|---|---|---|---|---|---|---|---|
| SARS-CoV-2 Antigen Rapid Test Kit (Colloidal Gold Immunochromatography) CE (pack of 25 tests) | 6921807601020 21CG2713X | 0,06 | 0,01 | 0,00 | 4-5 | 0,03 | 0,02 | 0,00 | 0,43 | 1,54 | n.d. |
| | 6921807601020 21CG2715X | 0,12 | 0,06 | 0,00 | 4 | 0,06 | 0,02 | 0,00 | 0,50 | 0,27 | n.d. |
| NASOCHECKcomfort SARS-CoV-2 Antigen Schnelltest Immunchromatographischer Test (kolloidales Gold) (pack of 25 tests) | CG2725(N) 21CG2722X | 0,06 | 0,04 | 0,00 | 2-3 | 0,04 | 0,04 | 0,00 | 0,73 | 1,04 | n.d. |
| NASOCHECKcomfort SARS-CoV-2 Antigen Schnelltest Zur Eigenanwendung (single pack; narrow) | CG2701N 21CG2727X | 0,10 | 0,04 | 0,00 | 2-3 | 0,07 | 0,02 | 0,00 | 0,69 | 0,38 | n.d. |
| NASOCHECKcomfort SARS-CoV-2 Antigen Schnelltest Zur Eigenanwendung (single pack; thick) | CG2701N 21CG2720X | 0,08 | 0,04 | 0,00 | 4-5 | 0,02 | 0,02 | 0,00 | 0,26 | 0,51 | n.d. |
| | CG2701N 21CG2724X | 0,13 | 0,04 | 0,00 | 4 | 0,07 | 0,01 | 0,00 | 0,52 | 0,37 | n.d. |

**Supplemental Figure S4: Variation in T/C ratios determined for different Lepu medical AgPOCT products.** (A) Means of T/C ratios (test band (T) intensity to control (C) band intensity) for each AgPOCT and Ct test sample (reproduced from Supplemental Figure S3). Numbers of replicates (n) are provided. (B) Standard deviations (SD) of T/C ratios for each Lepu medical AgPOCT product and Ct test sample. (C) Coefficients of variation (CV) of T/C ratios for each Lepu medical AgPOCT product and Ct test sample. Means, SD and CV are color-coded in shades of red (highest values with most intense color). Blue color highlights zeros. n.d. (grey) = not determined.
