## Supplemental Figure S5 for "Rapid comparative evaluation of SARS-CoV-2 rapid point-of-care antigen tests"

| | BfArM list? | BfArM ID | PEI evaluation? |
|---|---|---|---|
| **Watmind** | yes | 5640-S-032/21 | yes |
| **Clongene** | yes | 5640-S-168/21 | yes |
| **MP** | no | 5640-S-076/21 | yes |
| **Boson** | no | 5640-S-007/21 | yes |
| **Biomerica** | yes | AT007/21 | yes |
| **BTNX** | yes | AT102/21 | no |
| **Medice** | yes | 5640-S-128/21 | yes |
| **Testsea** | yes | AT082/20 | yes |
| **Beier** | yes | AT074/20 | yes |
| **Laihe** | yes | 5640-S-009/21 | yes |
| **ACON** | yes | AT261/21 | yes |
| **Healgen** | yes | AT079/21 | no |
| **möLab** | yes | AT047/20 | yes |
| **Teda** | yes | 5640-S-079/21 | yes |
| **Wondfo** | no | 5640-S-179/21 | yes |
| **Abbott** | yes | AT116/21 | yes |
| **nal von minden** | yes | 5640-S-045/21 | yes |
| **Realy** | yes | AT088/21 | no |
| **NanoRepro** | yes | 5640-S-096/21 | yes |
| **SD Biosensor** | no | 5640-S-025/21 | yes |
| **Deepblue** | no | 5640-S-086/21 | yes |
| **OFM** | yes | 5640-S-140/21 | yes |
| **Hotgen** | yes | 5640-S-057/21 | yes |
| **fameditec** | yes | 5640-S-154/21 | yes |
| **Sanicom** | yes | 5640-S-147/21 | yes |
| **RapiGEN** | no | ? | ? |
| **Safecare** | yes | AT376/21 | yes |
| **Joysbio** | yes | AT692/21 | no |
| **Ritter** | yes | AT527/20 | yes |
| **Joinstar** | no | ? | ? |
| **Hygisun** | yes | 5640-S-058/21 | yes |
| **Lepu Medical** | no | 5640-S-0104/21 | yes |

**Supplemental Figure S5: Information on BfArM listing and evaluation of investigated AgPOCTs by the Paul Ehrlich Institute** (as of July 27, 2021)**.** AgPOCTs are listed according to the ranking presented in Figure 1. (Middle column) AgPOCTs temporarily licensed for self-testing are provided with the respective BfArM GZ numbers (5640-S-XXX/20 or 21). AgPOCTs for professional use are listed with the corresponding BfArM test ID (ATXXX/ 20 or 21). (Left column) Presence of specified products (according to BfArM number given) on BfArM lists for rapid antigen tests for self testing or professional use (*) is indicated in light green. Note that if AgPOCTs for professional use are not BfArM-listed (light red; ? in middle and right column), they either failed PEI evaluation or manufacturers did not apply for BfArM listing. If AgPOCTs for self-testing are not BfArM-listed (light red), special permits might have expired or conformity assessments were completed. (Right column) Evaluation of AgPOCTs by PEI according to BfArM lists is indicated in light green. Note that AgPOCTs for self-testing with temporary special permits need a positive evaluation of the product for professional use. Light red indicates missing PEI evaluation.

*https://www.bfarm.de/DE/Medizinprodukte/Aufgaben/Spezialthemen/Antigentests/_node.html
