## Supplemental Table S6 for "Rapid comparative evaluation of SARS-CoV-2 rapid point-of-care antigen tests"

**Supplemental Table S6: Estimation of SARS-CoV2 genome copy numbers for Ct test samples.** Concentrations of the samples in SARS-CoV-2 genome copies per ml were calculated using internal data supplied by TIB MolBiol for the LightMix® Modular Sarbecovirus SARS-CoV-2 Kit, estimating 1000 viral genome copies per reaction at Ct30 and extrapolating therefrom. Numbers were corrected for the volume used in RT-qPCR (10µl) and the input/ eluate ratio of the RNA extraction process, and are given in logarithmic units base 10.

| Ct test sample | Copies per ml (corrected for extraction and RT-qPCR reaction volume) | log10 [SARS-CoV-2 genome copies/ ml] |
|---|---|---|
| 16 | 702 171 428 | 8,9 |
| 21 | 21 942 857 | 7,3 |
| 25 | 1 371 428 | 6,1 |
| 28 | 171 428 | 5,2 |
| Reference: 30 | 42 857 | 4,6 |
